# Association of DNA methylation age acceleration with digital clock drawing test performance: the Framingham Heart Study

**DOI:** 10.1101/2024.11.06.24316862

**Authors:** Zexu Li, Huitong Ding, Mengyao Wang, Yi Li, Ting Fang Alvin Ang, Gurnani Ashita, Gifford Katherine, Cody Karjadi, Daniel Levy, Rhoda Au, Chunyu Liu

## Abstract

**Background:** Cognitive function measured by digital clock drawing test (dCDT) has drawn attention for their precision, automation, and reproductivity. However, the relationship between digital cognitive metrics and biological aging is lacking.

**Methods:** We conducted association analyses between cognitive function measured by dCDT and biological aging metrics quantified by five DNA methylation (DNAm) age metrics (Horvath, Hannum, GrimAge, PhenoAge, and DunedinPACE) in the Framingham Heart Study (FHS). We conducted linear regression to investigate the association between cognitive functions (global cognitive function and four sub-domain functions) and DNAm age acceleration, adjusting for covariates. We used a false discovery rate (FDR) < 0.05 for significance.

**Results:** Among the 1,798 FHS participants (mean age 65±13, 53% women), we found that a lower dCDT total score is associated with DNAm age acceleration. Larger magnitudes of associations were observed in older participants (≥ 65 years). The dCDT total score showed the strongest association with the DundinPACE in the pooled sample (beta = −2.1, FDR = 0.0004), the younger (beta = −1.9, FDR = 0.02), and older age group (beta = −2.2, FDR = 0.01). The dCDT total score was significantly associated with age acceleration estimated by Horvath (beta=-1.9, FDR =0.01) and PhenoAge (beta=-2.5, FDR=0.01) in older participants while not in the pooled sample or younger participants (<65 years). In sub-domain cognitive functions, we found that simple motor function was significantly associated with DunedinPACE (FDR = 0.005) in both age groups and associated with GrimAge (FDR = 0.05 in older age group), indicating the deterioration in various organ systems may particularly impact this domain.

**Discussion:** Our findings suggest that cognitive function measured by a digital clock drawing test is associated with DNAm age acceleration in middle-aged and older participants in the FHS, potentially shedding light on the epigenetic mechanisms underlying digitally measured cognitive function.

## Introduction

Cognitive function, representing a crucial aspect of overall brain health, encompasses various domains such as memory, reasoning, and attention^1^. Neuropsychological (NP) tests are typically used to measure cognitive functions for individuals, focusing on one or several specific cognitive domains. For example, the traditional clock drawing test evaluates spatial dysfunction and neglect^2^, conducted with pen on paper. However, the assessment of the NP test is often biased. The use of computerized devices in NP tests is gaining recognition for their efficiency, precision, automation, and reproductivity nature^3^. The Digital Clock Drawing test (dCDT), an extension of the traditional clock drawing test, evaluates the overall cognitive function and specific sub-domains such as motor function, memory, spatial reasoning, and information processing^4^. Previous research has demonstrated strong associations of dCDT performance with mild cognitive impairment^5^, and with clinical indices of neurodegeneration, such as brain volume^6^ and NP tests^5^, showing a comparable performance between dCDT and other NP tests. Compared to traditional NP tests, dCDT can capture more subtle cognitive changes^7^.

Aging is an inevitable process for humans, resulting in a decline in physiological capacity and an increasing risk of various disease conditions^8^, including neurodegenerative diseases^9^. Biological aging, the changes at the molecular level^10,11^, is essential to understand the heterogeneity in healthy aging^12^. Epigenetic modifications, such as DNA methylation (DNAm), measure biological aging at the epigenetic level^13–15^. DNAm age^16^ has been associated with general health^17,18^ and neurodegenerative diseases^19^. The first generation of epigenetic clocks use methylation levels across varying numbers of 5’-C-phosphate-G-3’(CpG) sites with a few clinical markers to predict chronological age^16,20^. Subsequent generations of epigenetic clocks have incorporated additional clinical measurements to enhance their accuracy for age prediction^21–23^. DNAm age acceleration is assessed by regressing the estimated DNAm age on chronological age^24^.

Studies have established correlations between traditional cognitive assessments and DNAm age acceleration^25^, showing the possibility that DNAm age acceleration is a risk factor or marker for cognitive function decline. However, research on the relationship between cognitive function measured by digital devices and DNAm age acceleration is currently lacking. Our study aimed to fill in this gap. We hypothesize that cognitive function measured by digital devices is associated with DNAm aging. We investigated the associations of overall cognitive function and specific sub-domains measured by DCTclock with several DNAm-based age acceleration metrics in the Framingham Heart Study (FHS). We seek to gain additional insights into the underlying molecular mechanism of cognitive function measured by digital devices.

## Methods

### Study populations

FHS, initiated in 1948, is an epidemiological prospective cohort to study risk factors for CVD^26^. All FHS cohorts, including the original cohort (Gen1), offspring cohort (Gen2), and third-generation cohort (Gen3), have undergone routine health examinations every two to six years.

Our study included 1,264 Gen2 participants at exam 8 (2005-2008) and 688 Gen3 participants at exam 2 (2008-2011). We excluded participants whose blood samples were not collected at exam or who did not attend dCDT (2011-2018). After excluding participants with covariates (e.g., age, gender, education, cell counts) missing, we included 1,789 participants (**Figure 1**) in our statistical analyses. For participants with multiple dCDT tests, we selected dCDT data closest to DNA methylation measurement dates for inclusion in our study.

**Figure 1.**
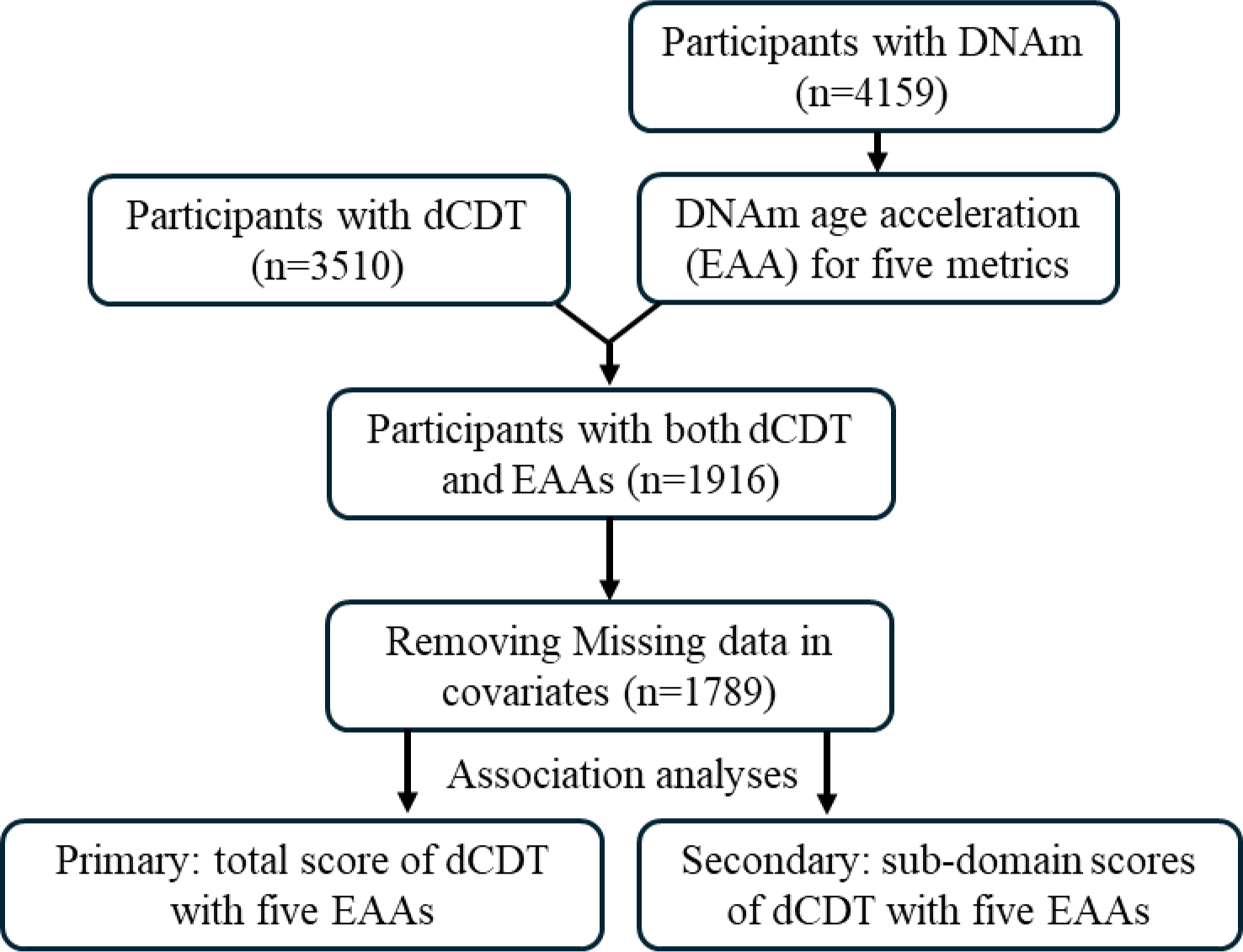
Flow chart of study design. The participation of dCDT was solely based on consent. In the Framingham Heart Study, we identified participants with digital clock drawing test (dCDT) measurements and DNA methylation (DNAm). Five DNAm age metrics were calculated. Epigenetic (DNAm) age acceleration (EAA) was calculated by regression of the DNAm age metrics on chronological age. Primary analysis focused on the association between dCDT total scores and EAAs, whereas secondary analysis focused on the association between dCDT sub-domain scores and EAAs.

### DCTclock

DCTclock serves as an FDA-approved automated screening tool for detecting cognitive changes^4^. Data collection for dCDT using DCTclock involved utilizing computerized neuropsychological assessment devices: digital pen and digital paper. Following the standard protocol, participants were instructed to draw clocks with the command ‘10 after 11’ and then replicated another clock by copying a provided model^27,28^. Both the drawing process and the final drawing results were recorded, capturing spatial and temporal data, and analyzed through the DCTclock pipeline. These data were treated as input to a trained convolutional neural network to recognize individual symbols with classification probability (e.g., clock face, digits, and small noise stocks). After classifying the individual symbols in drawing, these symbols were used to derive various measurements, such as the correct placement of clock components and pen speed. These measurements were organized into four groups representing different cognitive aspects: Drawing Efficiency, Simple and Complex Motor, Information Processing, and Spatial Reasoning. Drawing efficiency, for instance, evaluated the efficiency in terms of the time spent on drawing and the size of the drawing. Similarly, Simple and Complex Motor measurements represented motor and non-motor cognitive functions, including maximum movement speed. Information Processing focused on cognitive functions like thinking time and latencies, while Spatial Reasoning focused on spatial abilities through geometric property measurements. A composite score was calculated for each cognitive aspect mentioned above using a Lasso regularized logistic regression model^4^, incorporating all the measurements as parameters. Given the performance of two tasks (command and copy), eight sub-domain scores were generated. Additionally, a dCDT total score was computed using a Lasso logistic regression model^4^. Both sub-domain scores and the total score range from 0 to 100.

### DNAm measurements

DNAm adds a methyl group onto the 5th carbon of cytosine to form 5-methylcytosine^29^. DNAm measurements were conducted using whole blood samples collected during exam 8 for Gen2 and exam 2 for Gen3. DNAm profiling was carried out through a series of procedures, including bisulfite conversion, whole genome amplification, fragmentation, array hybridization, and single-base pair extension^30^. The Illumina Human Methylation 450K Bead chips (Illumina Inc., San Diego, CA) were employed to analyze the DNA methylation levels across three different laboratories. Detailed information regarding DNAm quantification and quality control procedures in FHS had been previously documented^31^.

### DNAm aging metrics

DNAm age is an estimator of aging based on DNAm patterns. Three generations of epigenetic clocks were calculated. The first generation, Horvath’s age^16^ and Hannum’s age^20^, utilize a set of CpG sites to estimate DNAm age. Horvath’s age calculated a weighted average of 353 clock CpGs with a calibration function to estimate aging from multiple tissues. Hannum’s age considered 71 CpG sites along with a few clinical parameters (gender, BMI, etc.) to predict aging in whole blood samples. GrimAge^22^ and PhenoAge^23^ are the second-generation DNAm aging metrics. Both methods calculated the DNAm age by integrating methylation levels with clinical markers. GrimAge first utilized DNAm data to estimate each plasma protein biomarker and smoking pack years. Then, 7 DNAm-based plasma protein biomarkers and DNAm-based smoking pack year were selected, which included 1,030 unique CpG sites along with gender and chronological age to predict time to death^22^. PhenoAge selected 513 CpGs to predict a linear combination of chronological age and nine clinical markers (e.g., Albumin, White blood cell count), which predicted the time to death^23^. We employed the principal component version of epigenetic clocks (PC-based clocks) to minimize unobserved technical confounders^32^. The DNAm age acceleration for the first- and second-generation aging metrics were residuals calculated by regressing each DNAm age on chronological age. Residuals larger than zero will be considered as accelerated aging. The third generation, DunedinPACE, differed from the previous generations by predicting the pace of aging per year rather than age in years^21^. The pace of aging was calculated from 173 CpGs based on longitudinal change of 19 clinical biomarkers (e.g., blood pressure, total cholesterol, blood urea nitrogen), representing an average rate of biological aging per year of 1-year chronological age^21^. This pace of aging was used as DNAm age acceleration for the following analysis.

### Covariates

Covariates included the age at the dCDT, the time interval between the dCDT and blood sample collection, gender, educational level, cell count information, and family relationship. The time interval was computed as the age at the dCDT minus the age at blood sample collection. Educational levels were categorized into four groups: less than high school completion, high school graduate, some college, and college graduate. Cell count information was derived from DNAm data. We included the count number of Cytotoxic T cells (CD8+T), B lymphocytes (CD19+ B), granulocytes (Gran), monocytes (Mono), Natural killer cells (NK), and Helper T Cells (CD4+T) as covariates. Family relationships were included as random effects in the model.

### Statistical analysis

The primary analysis explored the relationship between the dCDT total score as the outcome variable and various DNAm aging metrics as the predictor variables. The residuals were computed by regressing the DNAm age metrics on chronological age to obtain DNAm age acceleration. The residuals were used as DNAm age acceleration in the following analysis. To facilitate interpretation, we standardized the DNAm age residuals to have a mean of 0 with a standard deviation (SD) of 1. Participants were stratified into two age groups (<65 and ≥65 years), using age at blood sample collection, to address for age modification effects in the associations. Linear mixed models were employed to assess the association between the dCDT total score and DNAm aging metrics. We adjusted for age at the dCDT, gender, and educational level and used family as a random effect in both combined samples and age-stratified analyses.

To investigate the association between dCDT sub-domain scores and DNAm aging metrics, linear mixed models were employed with the same set of covariates. The False Discovery Rate (FDR) method^33^ was applied to adjust for multiple testing^34^.

## Result

### Participant characteristics

This study included 1,789 middle-aged and older participants in FHS (mean age 65 ±13 at dCDT, 53% women) (**Table 1**). On average, DNAm was measured seven years before the dCDT measurement (**Supplementary Figure 1**). Education levels were significantly higher in the younger age group (<65 years) compared to the older age group (≥65 years). On average, participants in the younger age group exhibited higher dCDT total score and sub-domain scores compared to those in the older age group (all p <0.001). The average estimated DNAm ages differed between DNAm metrics (**Supplemental Table 2**). For instance, the mean Hannum age was estimated at 62, while the Horvath age was estimated at 53 in pooled samples. Compared to the mean chronological age, the mean DNAm ages calculated by Hannum, GrimAge, and DunedinPACE showed acceleration (with DNAm age higher than chronological age) in the pooled sample, whereas PhenoAge and Horvath showed lower mean DNAm ages than the mean chronological age. We further observed that, except for DunedinPACE, males tended to have advanced DNAm ages compared to females (**Supplemental Table 3**).

**Table 1.**
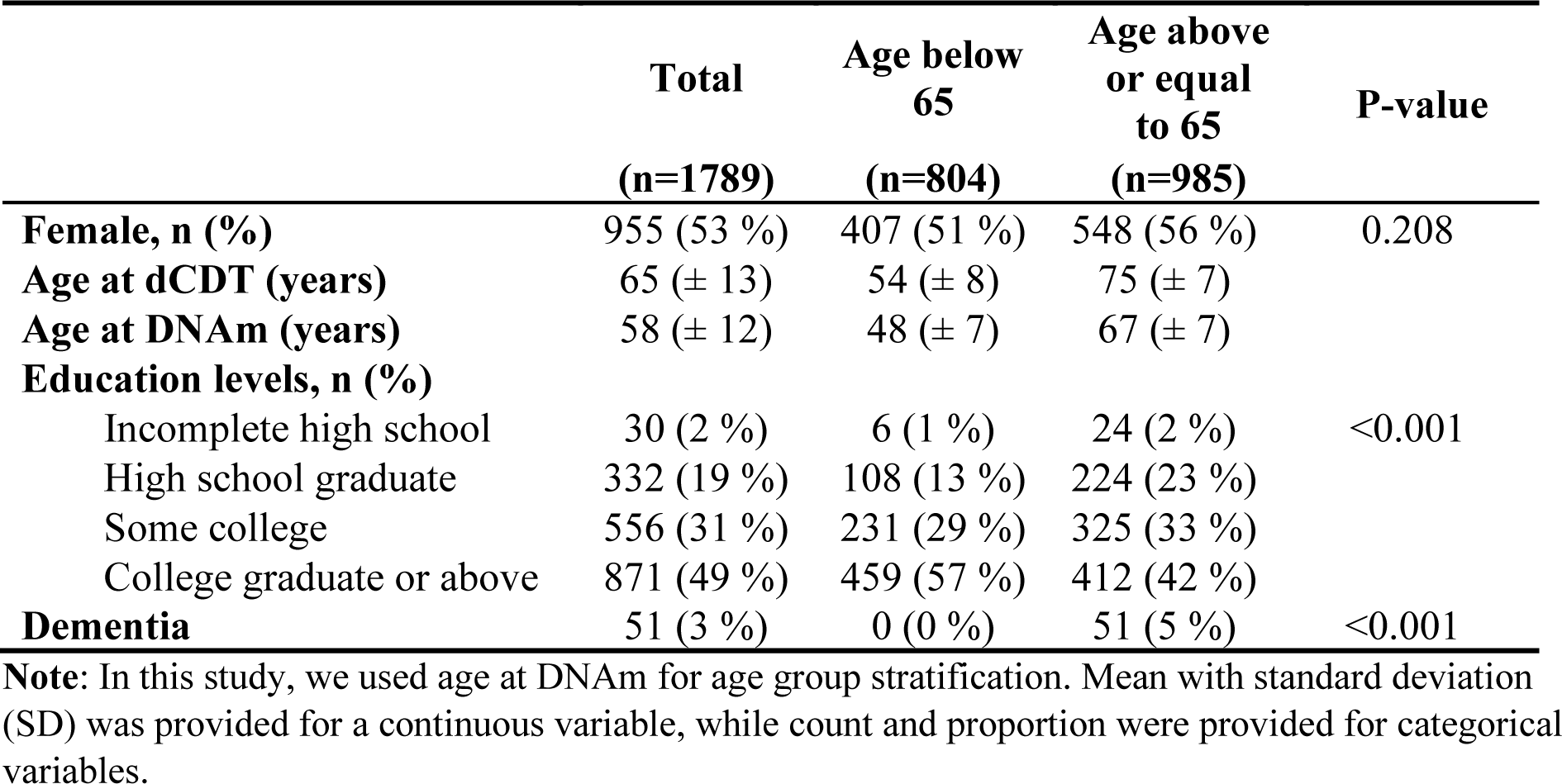
Demographic and Clinical characteristics of study participants.

### Association between dCDT total score and DNA methylation age acceleration

In the pooled sample, we observed that a higher dCDT total score was associated with lower DNAm age estimated by all DNAm aging metrics (**Figure 2**). However, the association was significant only for DunedinPACE, where a one-SD higher level in the pace of aging was associated with a 2.1-unit lower level in the dCDT total score (FDR=0.0004) (**Figure 2**).

**Figure 2.**
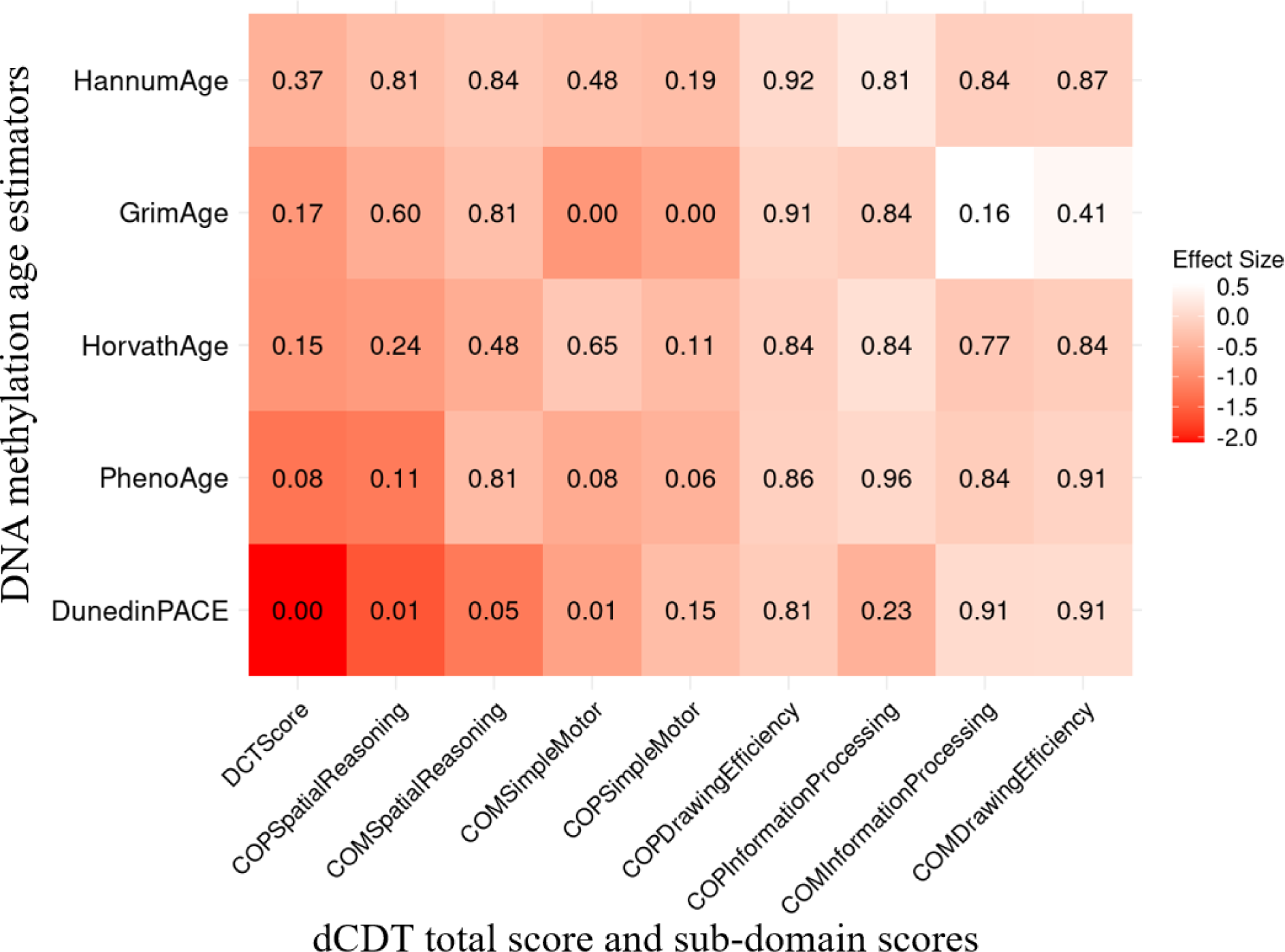
Association between dCDT scores and DNAm age acceleration in 1789 participants of the Framingham Heart Study. The dCDT total score includes command task composite scores and copy task composite scores. DNA methylation age acceleration was obtained by regressing DNAm age metrics on chronological age. We conducted association analysis between standardized DNAm age acceleration and the dCDT total score, adjusted for age, gender, and education. The numbers inside each cell represent the P-values of the associations. The color represents the change in dCDT scores corresponding to a one SD increase in DNAm age acceleration.

Age showed a significant effect modification of the association (p = 0.004) with DunedinPACE but not for other epigenetic age metrics (**Supplemental Table 4**). Thus, we conducted stratified analyses by age groups for all DNAm age metrics. In the younger age group (<65 years), DunedinPACE was the only DNAm metric associated with the dCDT total score (beta = −1.9, FDR = 0.02). In contrast, in the older age group, three DNAm aging metrics were significantly associated with the dCDT total score: DunedinPACE (beta=-2.2, FDR=0.01), DNAm age acceleration estimated by Horvath (beta=-1.9, FDR=0.01) and PhenoAge (beta=-2.5, FDR=0.01). Although the association between dCDT total score and other DNAm aging metrics was not significant, the directionality of the associations was consistent in both older and younger age groups (**Figure 3**, **Supplementary Figure 3**).

**Figure 3.**
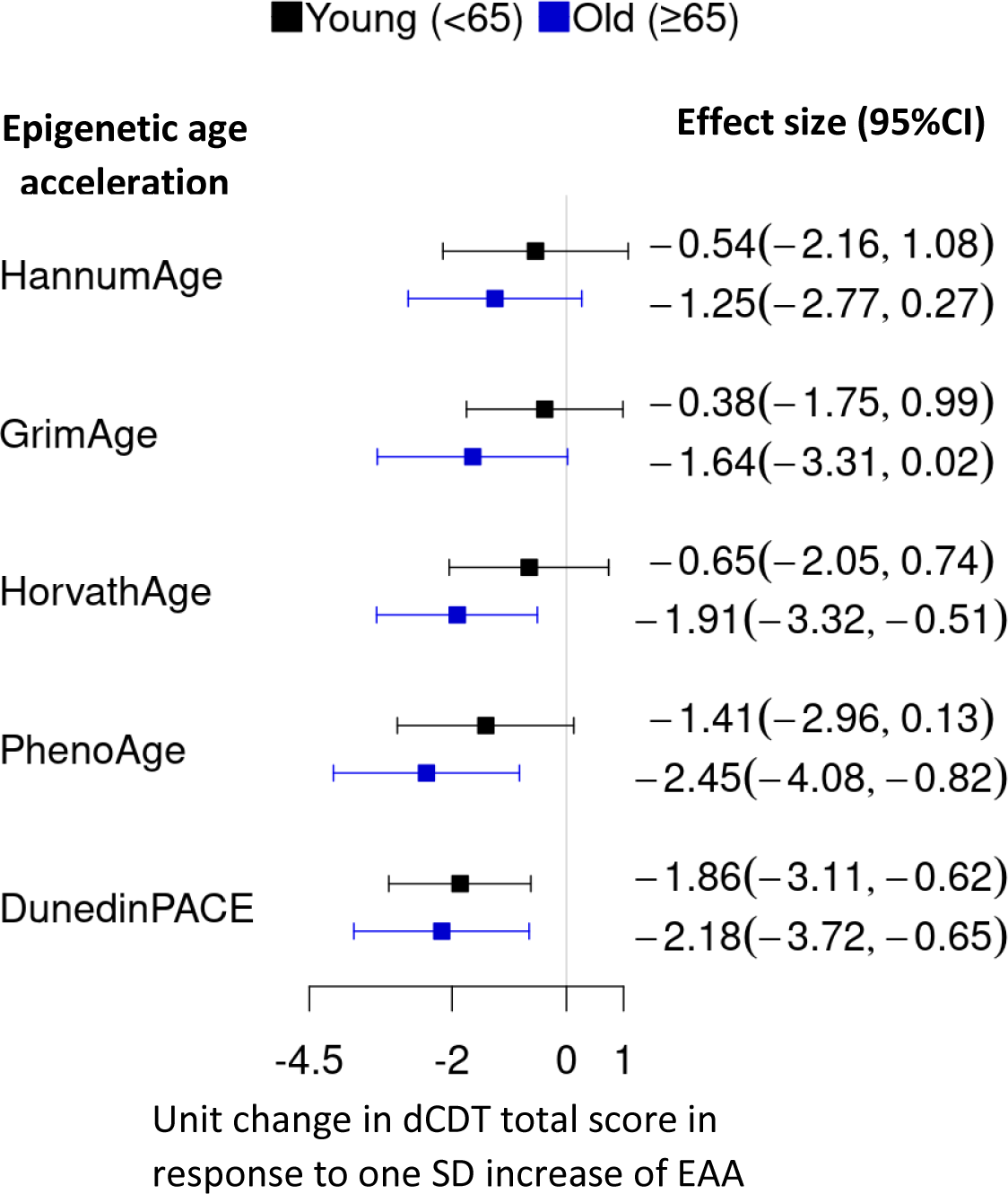
Comparison of effect size in the association between DNAm age acceleration and the dCDT total score. DNAm age acceleration was obtained by regressing DNA methylation (DNAm) metrics on chronological aging, followed by standardization with a mean of zero and SD of one. We conducted an association analysis between the dCDT total score and standardized DNAm age acceleration. CI, confidence interval.

### Association between dCDT sub-domain scores and DNA methylation age acceleration

In the pooled sample, we observed a significant association between DunedinPACE and both the dCDT simple motor function score in the command task (FDR = 0.01) and the spatial reasoning score from the copy task (FDR = 0.01). A one-SD higher level in the pace of aging was associated with a 0.7-unit decrease in the dCDT simple motor function score and a 1.6-unit decrease in the spatial reasoning score. GrimAge was also found to be significantly associated with the dCDT simple motor function score in both the copy task (beta = −0.7, FDR = 0.005) and command task (beta=-0.9, FDR = 0.005) in the pooled sample. No other significant association was found between the other epigenetic age acceleration metrics and the dCDT sub-domain score in the pooled sample (**Figure 2**).

Different from the result in the pooled sample, we only observed similar results for GrimAge in the older age group (>65 years), where a one-SD increase in GrimAge is significantly associated with a 1.1-unit decrease in dCDT simple motor function score in command task (FDR = 0.04) (**Supplementary Figure 2**). In addition, PhenoAge was significantly associated with the dCDT simple motor function score in the command task (beta =-1.0, FDR = 0.04). While no significant association was found between dCDT sub-domain scores and DNAm age in the younger age group, the directionality was consistent for most associations in both younger and older age groups (**Supplementary Figure 2, 3**).

## Discussion

In this study, we investigated the association between cognitive function measured by the dCDT and DNAm aging metrics in 1,789 middle-aged and older participants in the FHS. We found that lower dCDT total scores were consistently associated with advanced biological age quantified by DNAm aging metrics. Among all DNAm aging metrics, DunedinPACE showed significant association with the dCDT total score in the pooled sample, younger (<65 years), and older (≥ 65 years) age groups. In contrast, several other DNAm aging metrics showed significant associations with the dCDT total score only in the older participant group.

As the need for early diagnosis in the preclinical stage of AD grows, advanced screening tools for cognition are desired to capture subtle cognitive changes^35^. The dCDT has drawn attention for its efficiency, precision, automation, and reproductivity^3^. It has been demonstrated to be effective in distinguishing cognitive impairment from normal function^4,7^. Additionally, it is comparable to established NP tests, such as the Wechsler Memory Scale and Boston Naming Test, in discriminating between participants with mild cognitive impairment (MCI)^6^ and those with normal function. Unlike traditional paper and pen tests, the dCDT captures more granular data, including visuospatial, time-base, and kinematic details, offering a more detailed assessment. Its automated scoring system provides clinicians with objective and interpretable results^4^.

DNAm is a crucial epigenetic modification that influences gene expression without altering the underlying DNA sequence^29^. By incorporating different sets of CpG sites, the three generations of epigenetic age metrics offer varied perspectives on biological aging. Unlike the first- and second-generation clocks, which primarily estimate DNAm aging based on chronological age, clinical markers and time to death^16,20^, the third-generation clock, DunedinPACE, focuses on a set of different clinical markers (representing the progressive decline across multiple organ systems captured by multiple CpG sites)^21^. In our study, DunedinPACE is negatively associated with overall cognitive function in both younger and older age groups, while other age clocks only showed significant results in the older age group. These findings indicate that global cognition function, which is an indicator of brain health, may reflect the aging rates of multiple organ systems.

The analysis of dCDT sub-domain scores with DNAm age provides additional insights into how epigenetic aging may influence specific cognitive domains. For instance, the simple motor function and spatial reasoning sub-domain scores show significant associations with DunedinPACE, indicating that the deterioration in various organ systems may particularly impact these domains. GrimAge is based on seven plasma protein markers related to various diseases and conditions, including cardiovascular disease (Plasma B2M) and cognitive functions (Plasma B2M, ADM, cystatin C, and leptin)^16^. Our observation that simple motor function was associated with advanced GrimAge age indicates that the decline of simple motor functions might be related to abnormal levels of these protein markers in the plasma. Future studies are necessary to investigate the associations of these plasma proteins with cognitive decline.

Digital cognitive measures displayed stronger associations with most DNAm aging metrics among older (≥65 years) compared to younger (<65 years) participants, likely to reflect the cumulative and nonlinear age influences on both brain health and DNAm. This is consistent with our earlier findings of stronger associations between alcohol consumption and epigenetic age metrics^38^. For instance, overall cognitive function exhibited significant associations with PhenoAge in the older age group while not in the younger age group. In contrast, the global cognitive function score was associated with DunedinPACE in both age groups, indicating that DunedinPACE might be more sensitive to capturing the subtle influence of variations in the pace of aging on cognitive changes.

Several similar studies have investigated the association of cognitive function measured by traditional methods with DNAm aging metrics. Marioni et al.^39^ reported that general cognitive ability was associated with Horvath age in participants over the age of seventy. Our findings, using the dCDT, are consistent with this, showing similar associations with total score in older participants (65 years and above). Another study assessed how various cognitive tests (e.g., MMSE, ADAS-Cog-13, MoCA) associated with DNAm age acceleration in participants with a mean age of seventy-five^25^. They found that the faster pace of aging measured by DunedinPACE correlates with more severe cognitive decline^25^. Our results align with this, showing that worse cognitive function was associated with DunedinPACE, with a larger magnitude in older participants. Furthermore, our findings that higher PhenoAge and DunedinPACE were associated with poorer performance scores in older participants were also consistent with the previous findings, where they found that higher PhenoAge and DunedinPACE are associated with worse cognitive performance^25^.

Our study has limitations, including a lack of diversity (all participants were Whites) in our study sample. Further research with diverse groups is needed. In addition, there is an approximate 7-year gap between DNA methylation measurement and dCDT. This may introduce biases due to changes in DNA methylation levels and clinical conditions. Our study has several strengths. We employed the dCDT to measure cognitive function, which is a novel measure for assessing cognitive function. In addition, we employed PC-based clocks, which use principal components to reduce noise and enhance accuracy. To mitigate multiple testing, we applied FDR adjustment, which is more appropriate than Bonferroni correction for the presence of correlated outcome and predictor variables.

In conclusion, our study investigated how digital cognitive function assessed by dCDT relates to biological aging, measured by DNAm. These findings highlight the potential role of DNA methylation in cognitive function. Further research is needed to uncover the underlying biological pathways behind this association, particularly in more diverse populations.

## Data Availability

All data produced in the present study are available upon reasonable request to the authors and the Framingham Heart Study. The data used in this study could be requested through an application to the Framingham Heart Study (https://www.framinghamheartstudy.org/fhs-for-researchers/).

